# Spatial-temporal variations of atmospheric factors contribute to SARS-CoV-2 outbreak

**DOI:** 10.1101/2020.04.26.20080846

**Authors:** Raffaele Fronza, Marina Lusic, Manfred Schmidt, Bojana Lucic

## Abstract

The global outbreak of severe acute respiratory syndrome coronavirus 2 (SARS-CoV-2) infection causing coronavirus disease 2019 (COVID-19) reached over two million confirmed cases worldwide, and numbers are still growing at a fast rate. The majority of new infections are now being reported outside of China, where the outbreak officially originated in December 2019 in Wuhan. Despite the wide outbreak of the infection, a remarkable asymmetry is observed in the number of cases and in the distribution of the severity of the COVID-19 symptoms in patients with respect to the countries/regions. In the early stages of a new pathogen outbreak, it is critical to understand the dynamics of the infection transmission, in order to follow contagion over time and project the epidemiological situation in the near future. While it is possible to reason that observed variation in the number and severity of cases stem from the initial number of infected individuals, the difference in the testing policies and social aspects of community transmissions, the factors that could explain high discrepancy in areas with a similar level of healthcare still remain unknown. Here we introduce a binary classifier based on an artificial neural network that can help in explaining those differences and that can be used to support the design of containment policies. We propose that air pollutants, and specifically particulate matter (PM) 2.5 and ozone, are oppositely related with the SARS-CoV-2 infection frequency and could serve as surrogate markers to complement the infection outbreak anticipation.

## Introduction

The pandemic of the new coronavirus SARS-CoV-2 causing COVID-19 disease is testing the resilience of all the vital components of our communities, and specifically healthcare systems. SARS-CoV-2 belongs to a family of Coronaviridae, the common spillover pathogens that have recently caused two viral outbreaks, SARS and the Middle East respiratory syndrome (MERS). As SARS-CoV and MERS-CoV, new SARS-CoV-2 most probably originated from bats, however the intermediate host of the new coronavirus remains unknown to date (Wu et al. 2020). Like other coronaviruses, SARS-CoV-2 is a spherical enveloped virus with the positive-sense RNA genome. In general, coronaviruses have a relatively slow viral multiplication rate with the *in vitro* maximal replication efficiency at 32°-33°C, and a rapid decrease in infectivity at higher temperatures (Dulbecco and Ginsberg 1988). Moreover, coronaviruses can remain infectious for several days on inert surfaces and in the external environment (Firquet et al. 2015), (van Doremalen et al. 2020).

General indicator of transmissibility of a viral infection, basic reproduction number (R_0_), which indicates how many secondary infections are caused by a primary infected person, has been estimated for SARS-CoV-2 to be between 2 and 3 mostly using data from China (Liu et al. 2020). However, these estimations might not be accurate for what seems to be the faster-spreading European epidemic. The main transmission mode of SARS-CoV-2 is person-to-person contact (Ghinai et al. 2020). Besides physical contact, transmission can occur through physiological aerosol airborne droplets composed mainly of water in the act of breathing, talking, coughing and sneezing (Atkinsons J, et al. 2007), (Morawska and Cao 2020). The droplets that compose aerosol include various physiological constituents (cells, proteins, salts, small molecules) but also contain exogenous particles, such as viruses (Atkinsons J, et al. 2007). The dimension of these droplets varies, ranging from millimeters to microns, leading to a substantial difference in the detrimental effect in the case of infective particles (Stetzenbach, Buttner, and Cruz 2004), (Wong, Leung, and Fok 2004). Large droplets have a low probability to pass the upper respiratory tract, whereas the smaller particles have the capacity to reach the bronchi and lungs (Atkinsons J, et al. 2007). It is considered that droplets independently from the dimension, fall promptly to the ground as subordinated to gravity force, explaining the reason for the heuristic rule of maintaining a distance of a few meters to avoid the infection. However, the gravity force is counteracted by Stock’s friction and in the case of particles up to 4μm the two contrasting forces are equilibrating and the particles are floating in the air (Shaman and Kohn 2009).

Air pollutants, PM2.5 and PM10 are atmospheric aerosols that are classified on the basis of the particulate size (ie <2.5 μm and <10 μm). Toxicological studies suggest that PM2.5 are especially harmful as smaller particles are more prone to penetrate deeper into the lungs (Chan and Lippmann 1980). In line with this, several studies showed that exposure to particulate matter causes respiratory diseases (Xing et al. 2016), (Rajagopalan S. et 2018). Similarly, atmospheric pollutant ozone (O_3_), an oxidant present in the environment, also contributes to the risk of respiratory illness (Turner et al. 2016). However, unlike particulate matter, ozone is commonly used as a method for air, water and object disinfection. Indeed, in the case of an airborne influenza virus type A infection, O_3_ was found to rapidly inactivate the virus and to reduce morbidity in infected mice (Hiroshi et al., 2009) (Wolcott, Zee, and Osebold 1982), (Jakab and Bassett 1990).

In this study we analyzed particulate matter and ozone ambiental concentration in relation to SARS-CoV-2 infected cases from Italy and three other European countries, France, Germany and Spain. We propose that the same atmospheric conditions that elevate the air concentration of PM and O_3_, are also contributing to the modulation of the viral outbreak. We provide a qualitative model that predicts the outbreak severity taking into account only a reduced set of atmospheric factors. We propose that 1) atmospheric conditions that elevate the PM concentration are also involved in the SARS-CoV-2 viral outbreak, and that 2) the environmental ozone concentration can be a repressing factor that attenuates SARS-CoV-2 infection.

## Data and Methods

### Study setting and data

The hourly concentration (μg/m^3^) of PM2.5, PM10, O_3_ and NH_3_ in 53 regional capitals of Europe, and 107 major Italian cities, was retrieved in the period from 18^th^ February 2020 to 18^th^ of March 2020. For seasonal analysis, the first 30 days of March, June, September, and December were taken from the year 2018. The files are in crib2 or netCDF format retrieved from http://macc-raq-op.meteo.fr/. The analysis dataset used is the ENSEMBLE multi-model that combines the values of other seven models: CHIMERE METNorway, EMEP RIUUK, EURADIM KNMI/TNO, LOTOS-EUROS SMHI, MATCH FMI, SILAM Météo-France, and MOCAGE UKMET (https://www.regional.atmosphere.copernicus.eu). The European domain is defined within the coordinates −25E/70N and 45E/30N, with 0.1 degrees of horizontal and vertical resolution. The values were extracted and manipulated into a Linux environment using bash scripts, cdo 1.7.0 (DOI 10.5281/zenodo.1435454), and the R packages raster (Robert J. Hijmans, 2020) and ncdf4 (David Pierce, 2019).

The concentration of PM2.5, PM10, O_3_, and NH_3_ are expressed in μg/m^3^ within an area of about 64km^2^ where the geographical centre of the province capital corresponds to the geometric center of the quadrant. The latitude and longitude of Italian cities were derived directly from the epidemiological data provided by the Protezione Civile. The latitude and longitude of European cities were obtained manually.

Inside the Italian province dataset we subdivided the data in three macroregion: north Italian provinces (latitude ≥ 44.85), central Italian provinces (41.5 ≤ latitude < 44.85), and south Italian provinces (latitude < 41.5).

To express the overall quantity in the study period for PM2.5, PM10, NH_3_, and O_3_, we developed and tested two indexes, the average daily maximum (AdM) and the average daily value (AdV). Considering the starting day of the atmospheric factors sampling (02.22.2020), the incubation period (6 days, (Backer, Klinkenberg, and Wallinga 2020)) and the day at which we started the survey (03.25.2020.), D was imposed as *D* = 27. Considering that there are *L* geographical areas, and 24 hours a day (*H*), the *AdM* at a day *d* is a vector computed as:

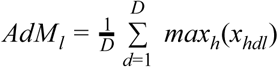

where *x_hdc_* is the meteorological factor value at hour *h*, day *d*, and location *l*. The *AdV* is computed as:

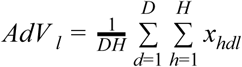

The two indexes are similar with *AdM* being more sensitive to rapid changes of the factors whereas *AdV* being more robust to spikes and responses to temporal consistent changes. For ozone, the concentrations were converted from μg/m^3^ to ppm assuming standard conditions (273.1° *K*, 1 atm) and using the relation:

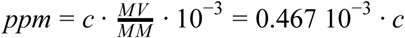

where *c* is the ozone concentration (μ*g*/*m*^3^), *MV* is the molar volume approximated with an ideal gas (22.414 l/mol) and *MM* is the molar mass (48 g/mol).

For each species, we obtained a 1xn vector at a final day *D*, where *n* is the number of areas considered.

### Epidemiological data

Given the open data policy of the Italian Government, for Italy it was possible to obtain the daily data of the SARS-CoV-2 infected people per province and region. The demographic data were collected from the site http://www.comuni-italiani.it/provincep.html. The Italian SARS-CoV-2 case data are retrieved from https://github.com/pcm-dpc/COVID-19 in the form of daily comma-separated files. For each day we collected a set of *m* × *n* matrices where m is 31, the number of days (from 24^th^ of February to 25^th^ of March) and n is the number of the provinces or regions.

In the case of the other three European countries, France, Germany and Spain (FGS), the data were not centralized or specified at the time of the analysis and were manually imported without the possibility to separate in subgroups of cases. The total number of cases for 18 French, 16 German and 19 Spanish regions, were obtained from https://www.statista.com/ referring to 25^th^ of March.

### Statistical analysis

The pairwise Spearman Correlation Coefficients were computed for all the atmospheric factors. Conditioning plots were then applied to graphically assess the dependence of the number of infected cases per million of one factor conditioned to different levels of another factor.

A generalized Poisson model was fitted to estimate the association among the data showing the number of infected cases per million and the atmospheric factors. The infection data used per province are the total number of patients on 25^th^ of March, whereas for the regions we used the number of total, not hospitalized and hospitalized cases. We calculated *AdM_l_*, where *l=* PM2.5, PM10, O_3_ and NH_3_ considering D=27 (endpoint 19^th^ March). All the regression models were tested and adjusted for overdispersion.

### Binary classifier

Binary classifier based on an artificial neural network (ANN) was implemented to test the capacity of the atmospheric variables to predict the epidemic escalation of the number of positive cases per million on the basis of a combination of *AdM_l_* where *l=* PM2.5, PM10, NH_3_ and O_3_. All the *AdM_l_* values were pre-scaled to be homogeneous. We defined as epidemic escalation, when the number of cases per million (at the time of the present analysis) is more than one standard deviation (σ = 480), above the mean (μ = 543) of the number of cases per million in the first twenty nations listed in the coronavirus survey https://www.worldometers.info/coronavirus/. We call this value escalation threshold (*T_e_* = 1023 cases per million). The binary classifier was developed using a feed-forward ANN with n inputs, one output and h hidden components using the *neuralnet* R package. The number of inputs n depends on the number of regressors used as input, whereas the number of the hidden components was selected during the training phase with the Italian province datasets. The dimensionality of the input parameters was finally reduced to the usage of two variables, PM2.5 and O_3_.

To validate the models we selected a Monte Carlo cross-validation strategy. The observed number of data was split randomly multiple times (n=100) into two datasets, the training (*R*) and the test (*T*), containing 70 and 37 provinces respectively. For each combination of regressors, we varied the number of hidden layers from 3 to 15. We defined as true negative (*TN*) all the provinces in T that have a number of cases that is lower than the selected threshold. The true positives (*TP*) are provinces where the number of cases is higher than the threshold. The false positives (*FP*) are all the provinces predicted with epidemic escalation but where the number of cases is lower than the threshold. The false negatives (*FN*) are all the provinces predicted without epidemic escalation but where the number of cases is higher than the thresholds. The random assignment to *R* or *T* was performed 100 times for each combination of the parameters. A null predictor was implemented with a random swapping of the number of cases. To measure the performances of the binary classifier and the null predictor, we used four indices: the sensitivity 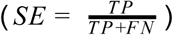, the specificity 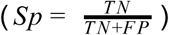, the accuracy 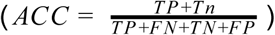, and the precision 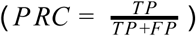 for PM2.5, PM10, NH_3_, O_3_, and the combinations of the four factors. The best model and threshold that maximizes the sum of all four indices on an average of 100 training-testing phases were selected.

### Classifier evaluation

To evaluate the capacity of the classifier we collected the total number of SARS-CoV-2 cases for three European nations, France, Germany and Spain from 25^th^ of March. The atmospheric factors PM2.5 and O_3_ were extracted in the same way as described in the Study setting and data section. The data were then scaled following the same procedure, and the conditioned factors were used to feed the classifier to measure the performances on completely unseen data. The expected number of infected cases in the total of 107 Italian provinces were predicted for the months of March (Spring), June (Summer), September (Autumn) and December (Winter) using the real measured values for PM2.5 and O_3_ atmospheric factors from 2018 seasonal datasets.

### Informal Boolean approximation

The results of the predictions and the previous results were condensed into a simplified boolean model. The explaining continuous factors, PM2.5 (*V*(*p*)), and ozone (*V*(*o*)), where *p* and *o* are concentrations, are seen as boolean variables that refer to two states, present (*V*(*p* > *T_p_*), *V*(*o* > *T_o_*) or not present, depending on the unknown thresholds (*T_p_*, *T_o_*). As the atmospheric factors considered are limited source that influences the severity of infection, all the hidden factors are summarized by a single hidden variable *V*(*u*) with unknown characteristics. The effect of the combination of the three variables on the epidemic has two states, escalating and non escalating, on the basis of the described epidemic escalation threshold *T_e_*. This model is qualitative, and the activation thresholds *T_p,o_* are not explicitly calculated. The observation that the modality of the interactions among factors derived from the regression step is not known is justifying the usage of an ANN as a general approximator.

## Results

The concentration of four air pollutants in 107 Italian and 47 FGS areas were sampled over a period of 27 days from 00 AM on the 21^st^ of February until the 00 AM of the 18^th^ of March. The average population of an Italian province is 572,646 inhabitants with an average number of cases per million of 1226 at the time of the study. On the same day the average population and number of cases per million in the FGS regions was 4,241,214 and 1,097, respectively. In the first 20 countries, ranked by the total number of cases at the day of the study, the average number of cases per million was 480.

To measure the strength of association between all the variables in the provinces we used the Spearman coefficients (SCs) (Table 1). All the *AdM_PM2.5_*, *AdM_PM_*_10_, and 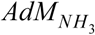 values were nearly identical with the respective *AdV_l_* values (r>0.99) suggesting that the two indexes are redundant. The 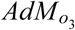 and 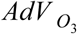 values were correlated with a slightly lower SC (r=0.91). Correlation matrix for PM2.5, PM10 and NH_3_ shows a strong positive inter-variables correlation (r>0.7). The number of SARS-CoV-2 cases per million (Cas) shows a significant positive correlation with three of the four atmospheric variables PM2.5, PM10, and NH_3_ (0.58 ≤ *r* ≤ 0.68). O_3_ instead, shows a good negative correlation with Cas (−0.5 ≤ *r* ≤−0.44). We also introduced the total population of the province (Pop) to assess a correlation between the number of inhabitants and atmospheric pollution. A weak correlation was found between the population, PM2.5 and PM10 (0.26=<r<=0.31). Moreover, no or little correlation was found among the population, NH_3_ and O_3_. Overall, SCs show that all the tested atmospheric variables display an extremely high degree of intra-variable correlation, allowing us to half the dimensionality of the variable space. High inter-variable correlation among PM2.5, PM10, and NH_3_ makes these variables redundant.

**Table 1.** The correlation matrix between PM2.5, PM10, NH3, O_3_, population and number of SARS-CoV-2 cases in Italian provinces. Suffix s in the columns and rows names (e.g. PM10s) corresponds to the *AdV (AdV_PM_*_10_) whereas the suffix m (e.g. PM10m), represents *AdM* (AdM_*PM*10_).

| Spearman Correlation Coefficients |  |  |  |  |  |  |  |  |  |  |
| --- | --- | --- | --- | --- | --- | --- | --- | --- | --- | --- |
|  | Pop | PM10m | PM10s | PM2.5m | PM2.5s | NH<sub>3</sub>s | NH<sub>3</sub>m | O<sub>3</sub>m | O<sub>3</sub>s | Cas |
| Pop | 1,00 | 0,31 | 0,30 | 0,28 | 0,26 | 0,07 | 0,08 | -0,18 | -0,29 | -0,03 |
| PM10s | 0,31 | 1,00 | 0,99 | 0,99 | 0,98 | 0,76 | 0,79 | -0,59 | -0,79 | 0,59 |
| PM10m | 0,30 | 0,99 | 1,00 | 0,97 | 0,98 | 0,79 | 0,81 | -0,56 | -0,77 | 0,58 |
| PM2.5m | 0,28 | 0,99 | 0,97 | 1,00 | 0,99 | 0,76 | 0,78 | -0,62 | -0,82 | 0,60 |
| PM2.5s | 0,26 | 0,98 | 0,98 | 0,99 | 1,00 | 0,80 | 0,81 | -0,61 | -0,82 | 0,60 |
| NH<sub>3</sub>s | 0,07 | 0,76 | 0,79 | 0,76 | 0,80 | 1,00 | 1,00 | -0,47 | -0,62 | 0,68 |
| NH<sub>3</sub>m | 0,08 | 0,79 | 0,81 | 0,78 | 0,81 | 1,00 | 1,00 | -0,47 | -0,63 | 0,69 |
| O<sub>3</sub>m | -0,18 | -0,59 | -0,56 | -0,62 | -0,61 | -0,47 | -0,47 | 1,00 | 0,91 | -0,44 |
| O<sub>3</sub>s | -0,29 | -0,79 | -0,77 | -0,82 | -0,82 | -0,62 | -0,63 | 0,91 | 1,00 | -0,50 |
| Cas | -0,03 | 0,59 | 0,58 | 0,60 | 0,60 | 0,68 | 0,69 | -0,44 | -0,50 | 1,00 |

To select the variables that mostly contribute to explain the escalation in the number of cases, we applied the generalized Poisson model with the log-transformed infected counts. We reduced the number of variables selecting only the interactions among the *AdM* atmospheric factors, excluding the *AdV* as redundant. Hence, from now on we refer to the *AdM_l_* index only, so that the factor species name *l*, as for example PM2.5, will be used throughout the manuscript. The latitude of the province capital was introduced as an explaining factor in the regression model. The location of the province in respect to the latitude was found as a major factor (p<0.004279) with a positive trend that interacts significantly with PM10 and O_3_. The amount of PM, in the south of Italy (green dots, lat<41.5), occupy the left part of the scatter plot, remaining under 30μg/m^3^ (PM2.5) and 40μg/m^3^ (PM10) (Figure 1A and B). The rest of the provinces show a more extended range of values. The distribution of O_3_ (Figure 1C), is instead more uniform in a range that goes from 0.03 to 0.05 ppm. In the south Italian provinces, this factor shows no association with the total number of cases. In central Italy (41.5<lat<44.85) and in northern Italy (lat>44.85) the tendency of the O_3_ concentration has clearly a negative relation with the number of SARS-CoV-2 positive cases. The validity of the regression model passed the check for normality, heteroscedasticity, influence and scale-location (Supplementary Figure S1). To correct for overdispersion we removed systematically all the explaining variables from the model and performed χ^2^ test to assess the significance of the deleted variables. As expected after the correlation analysis, PM2.5, PM10 and NH_3_ were strongly positively correlated (p<0.001) with the number of cases whereas O_3_ showed a significant (p=0.013) inverse correlation.

**Figure 1.**
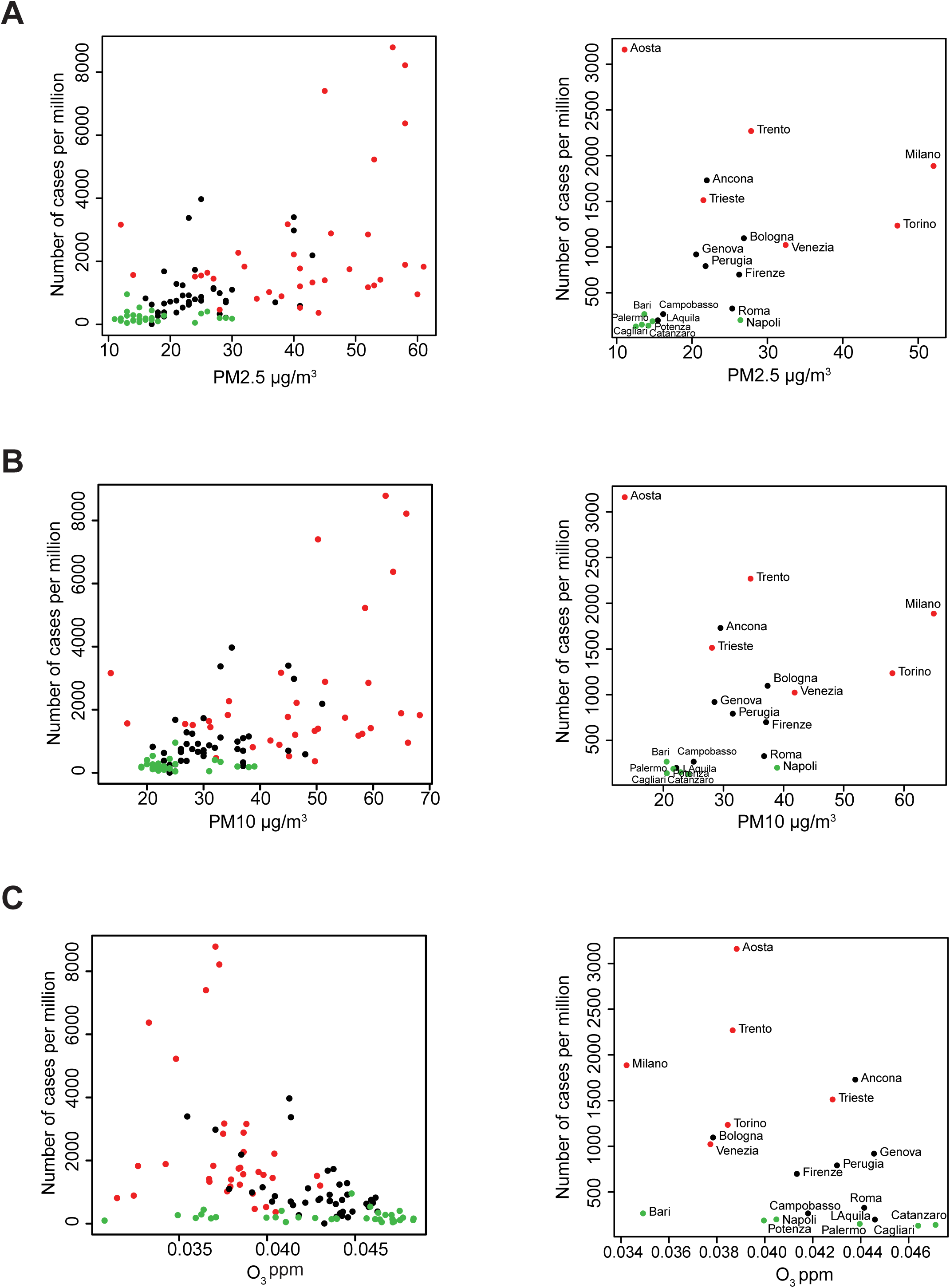
Correlation between SARS-CoV-2 cases in 107 Italian provinces using PM2.5, PM10 and O_3_. The scatterplots display the values of three atmospheric factors and the number of cases per million in 107 provinces (left panels) and selected provinces with regional capitals (right panels). A) PM2.5; B) PM10; C) O_3_. Different colours represent Italian provinces at different latitudes. Red dots: provinces with a latitude bigger than 44.84N; black dots: provinces with a latitude comprise between 41.50N and 44.86N; green dots: provinces with a latitude lower than 41.50N.

To visually detect relations that may be obscured by the effects of other variables, a collection of conditional plots (Cleveland 1994) was generated (Figure 2 and Supplementary Figure S2). We analyzed PM2.5 (Figure 2A) and the O_3_ (Figure 2B and 2C) as the explanatory variables for the number of cases and PM2.5 (figure 2A and 2C) and O_3_ (Figure 2B) as conditional values. When the variables are self-conditioning (Figure 2A and 2B), the number of SARS-CoV-2 cases per million depends on PM2.5 and O_3_ on an apparent opposite ways; PM2.5 at concentrations higher than 30 μg/m^3^ (fifth and sixth scatter plot in figure 2A) and O_3_ at concentrations lower than around 0.04 ppm (first and second scatter plot in Figure 2B). The number of cases per million starts to be dependent on O_3_ concentration only when the PM2.5 are higher than about 30μg/m^3^ (fifth and sixth scatter plot in Figure 2C). In summary, conditional plots show that the number of cases per million appears to be non linearly linked with the O_3_ data due to a threshold effect on the ozone linked to the PM2.5 concentration. The same threshold effect seems to hold true for the other two analyzed variables, PM10 and NH_3_ (Supplementary Figure S2).

**Figure 2.**
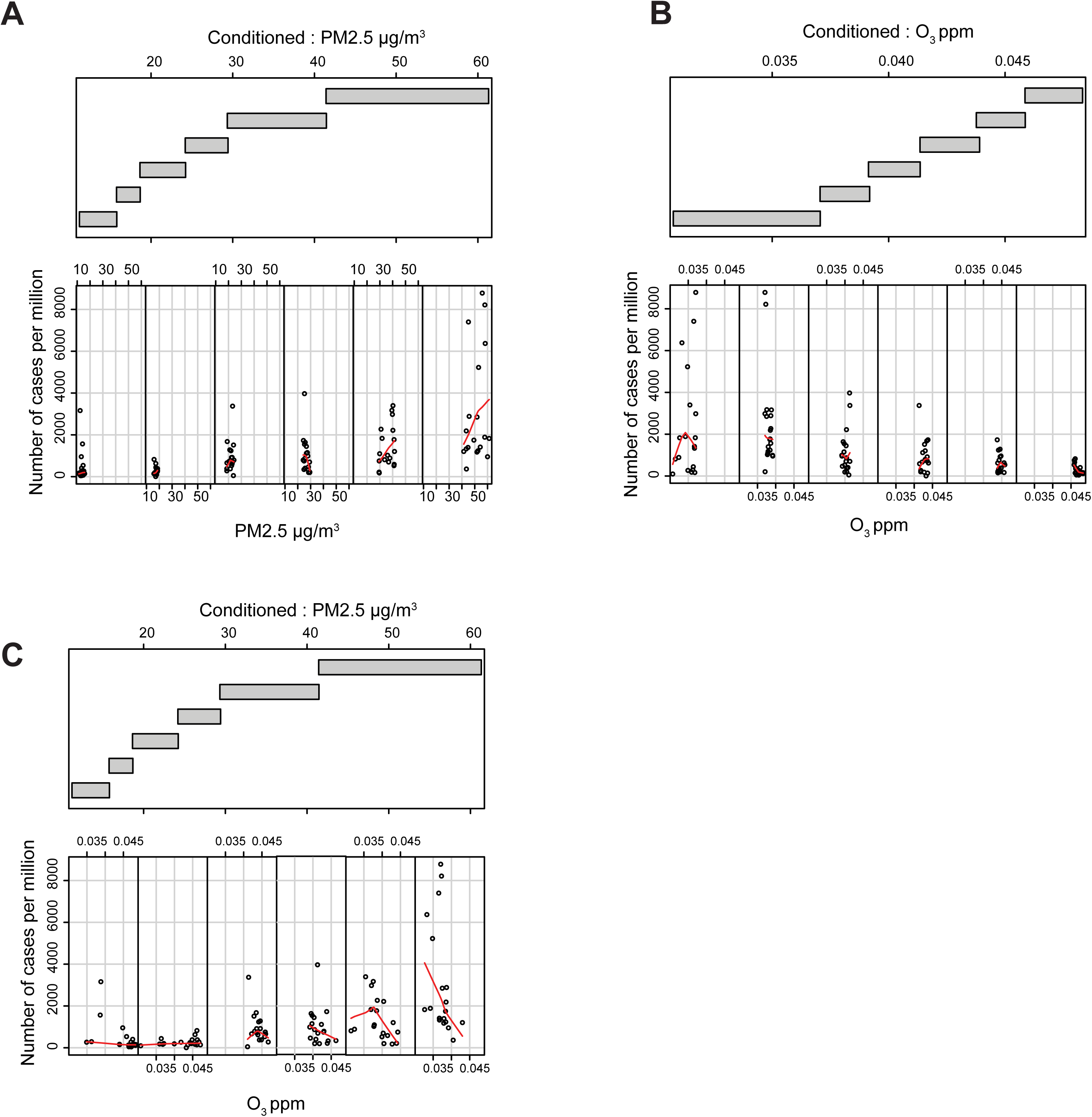
Evaluation of the cross-influence between PM2.5 and O_3_. Lower panel: the scatter plots between the selected factor and the number of cases per million. Each plot is restricted to show data points that belong to the provinces that fall in the corresponding range of the conditioning factor. Upper plot: the range of the values that define each level of conditioning. The overlapping among the levels is 0.1. The lowess curve is computed in each scatterplot and shown in red. A) the scatterplot of O_3_ conditioned to O_3_; B) the scatterplot of PM2.5 conditioned to PM2.5; C) the scatterplot of O_3_ conditioned to PM2.5.

The ANN classifier with three hidden neurons (Figure 3A) returned the highest score in SE, SP, ACC and PRC (Figure 4A and Supplementary Figure S3A), so this topology was selected to classify the datasets.

**Figure 3.**
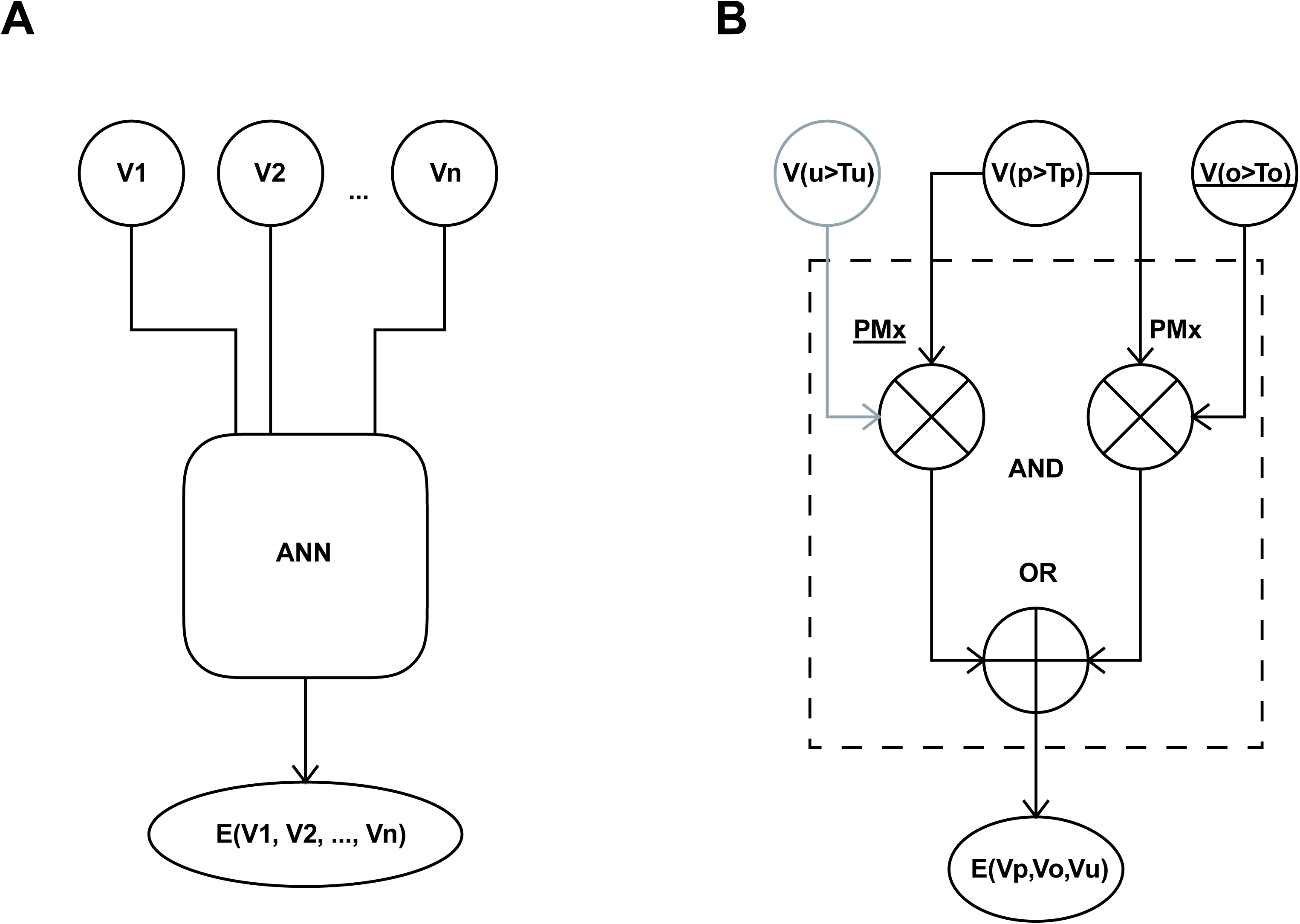
ANN and approximated logic model schema. A) The schematic design of the AAN classifier. A set of N input nodes are filled with a combination of the measurement of atmospheric factors V_1_, V_2_, …, V_n_. The output of the ANN is a function E(V_1_, V_2_, …, V_n_) on the atmospheric variables V_1_, V_2_, …, V_n_. B) Intuitive boolean diagram of the model suggested by our conclusions. V_p_, V_o_, and V_u_ are three input signals that correspond to PM2.5, O_3_, and an unknown generic variable. The three variables are mapped to {0,1} using a threshold function where *V_x_* = 1 if the concentration of x is bigger than a threshold T_x_ and *V_x_* = 0 otherwise. The threshold T_x_ is implicitly determined by the neural network. The notation 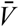 indicates the negation of *V*. ⊕represents an OR and ⊗ an AND gate. *E(V_p_, V_0_, V_u_)* is the binary output of our model given V_p_, V_o_, and V_u_.

**Figure 4.**
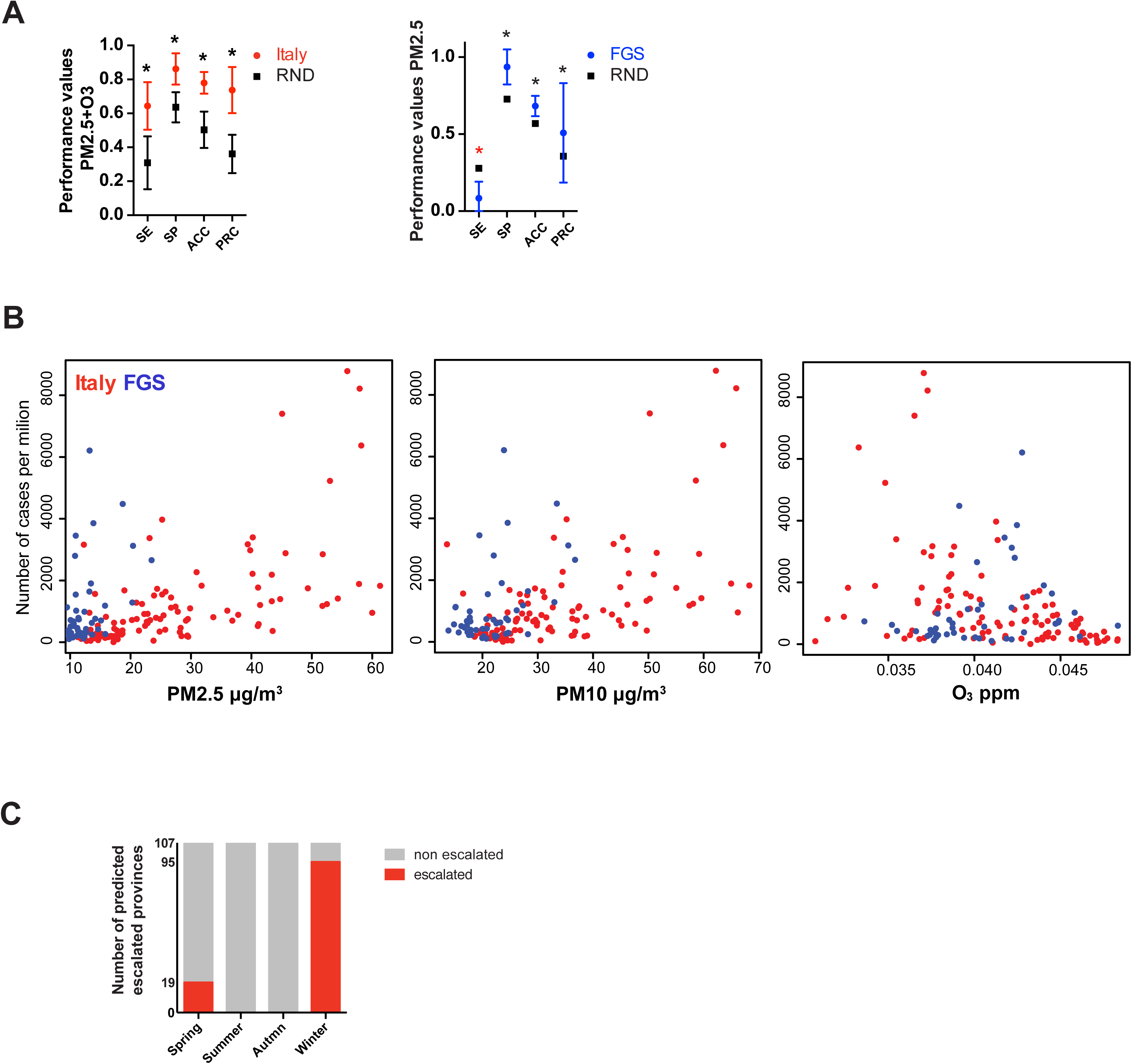
ANN performance assessment. A) Performance values of the ANN on the 107 Italian provinces data. SE sensitivity, SP specificity, ACC accuracy, PRC precision. The dots represent the ANN average performance based on 100 Monte Carlo cross-validations. Bars represent the standard deviation. Red dots Italian provinces, blue dots FGS regions and black dots Random dataset. B) The scatterplots display the concentration of PM2.5, PM10, and O_3_. Red dots Italian provinces, blue dots FGS regions. C) The histogram represents the number of the escalated Italian provinces (107) considering the PM2.5 and O_3_ concentrations in four months, March, June, September, and December, representative of the four seasons: Spring, Summer, Autumn, and Winter. Red bars, number of escalated provinces, grey bars, remaining non escalated provinces. Statistical analysis were performed using multiple t test corrected with Sidak-Boneferroni method for multiple comparisons (* p-value<0.001). Black asterisk indicates that the classifier performs better than the null classifier. Red asterisk indicates that the classifier performs worse than the null classifier.

The prediction on the Italian provinces using PM2.5, O_3_ and both variables are summarized in Supplementary Figure S3B and Figure 4A, left panel. The classifier with both PM2.5 and O_3_ has sensitivity, specificity, accuracy, and precision that are highly significant in respect to the null predictor. The usage of only PM2.5 or O_3_ is still significant but with a decreased capacity to predict the escalated provinces (Supplementary Figure S3B).

For the FGS data the results show a lower capacity of the combination of PM2.5 and O_3_ in classifying performances (Supplementary Figure S3A). When only PM2.5 was used (Figure 4A, right panel), the predictor behaved better than the null model for specificity, accuracy and precision. Remarkably, the classification efficiency of O_3_ alone was sub-performing for all the four performance indexes. When we combined the data from Italian provinces and FGS regions, the distribution of the PM (Figure 4B, left and middle panels) in the FGS regions was clearly lower, similar to the value observed in the south of Italy (<30μg/m^3^). The distribution of the ozone concentration is narrower in the FGS regions, with a shorter tail at the lower concentrations (Figure 4B, right panel).

Next, we tested the classifier seasonal predictive behaviour taking the *AvM* from the 107 Italian provinces, extracted from the seasonal recordings of March (Spring), June (Summer), September (Autumn), and December (Winter) 2018 (Figure 4C). Zero outbreaks were predicted in June and September (Summer and Autumn), whereas 19/107 outbreaks were found in March (Spring) and 95/107 in December (Winter).

Taken together, classification results show that the low concentration of the particulate is a reliable predictor for the absence of epidemic escalation. At medium-high values, the model predicts well the epidemic escalation in combination with low (high epidemic escalation likelihood), or high (low epidemic escalation likelihood) concentration of ozone. This observation makes the ozone a possible predictor only if the quantity of particulate is significantly high. For all the conditions where the PM2.5 is low, the unknown factors explained by unknown variables (V_U_) are prevailing in explaining the epidemic escalation.

Finally, we explore the hypothesis that atmospheric conditions that favor the formation of PM and possibly viral micro droplets differentially influence the severity of SARS-CoV-2 infection. By separating hospitalized and not hospitalized cases in the 21 Italian regions, we found a significant positive correlation (R^2^=0.4891, p=0.0004) between the particulate matter quantities and the number of hospitalized patients (Figure 5A), whereas for infected but non hospitalized population, this was not the case (R^2^=0.01154, p=0.6430). Ozone instead, showed a significant negative correlation with the hospitalized population (R^2^=0.2355, p=0.0257) and a not significant negative correlation (R^2^=0.1590, p=0.0734) with the not hospitalized population (Figure 5B).

**Figure 5.**
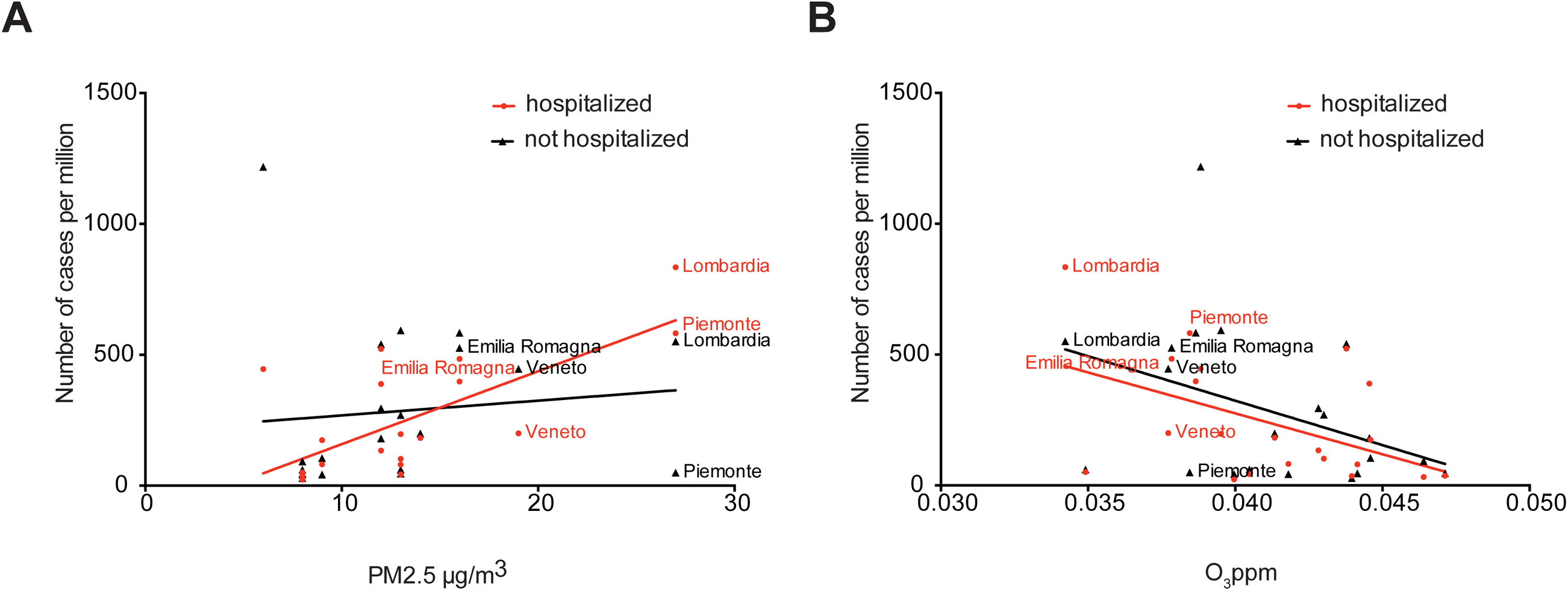
Correlation between hospitalized and not hospitalized cases in 21 Italian regions using PM2.5 and O_3_. A) Scatter plot of the hospitalized (red points) and not hospitalized (black points) versus the concentration of PM2.5 in 21 Italian regions. Red line, positive correlation (R^2^=0.4891, p=0.0004); black line, no significant correlation (R^2^=0.01154, p=0.6430). The labels represent the four regions with the highest number of cases per million. B) Scatter plot of the hospitalized (red points) and not hospitalized (black points) versus the concentration of ozone in 21 Italian regions. Red line, negative correlation (R^2^=0.2355, p=0.0257); black line negative correlation (R^2^=0.1590, p=0.0734). The labels represent the four regions with the highest number of cases per million.

To continuously monitor the capacity of our framework to predict the spread of the SARS-CoV-2 in the 154 areas, we regularly update the PM2.5 and O_3_ data and perform a real-time forecasting of the infection. The results of this analysis is provided as a pdf document in https://github.com/COVID19Upcome/Europe.

## Discussion

In the present study, we built a predictive model for the SARS-CoV-2 viral outbreak as a function of atmospheric pollutants, PM2.5, PM10, NH_3_ and O_3_. Our hypothesis is based on the correlation between viral outbreak and physicochemical factors that contribute to the enrichment of the atmospheric particulate.

Based on the Spearman, conditional and regression correlation analysis, we chose to favor the use of the average daily maximum values of PM2.5 and O_3_ to build the ANN classifier for prediction of SARS-CoV-2 outbreaks. The ANN model with two input variables, PM2.5 and O_3_, and three hidden layers was trained with 107 Italian provinces, which resulted in a significant predictive ability for all four considered performance values (Figure 4A and Supplementary Figure S3B). The same model however, tested on FGS regions, showed a limited capacity to classify the epidemic escalation in those regions. A more in-depth analysis taking into account predictors with only a single atmospheric input variable resulted in the amelioration of the predictive capacity with only PM2.5 in the FGS regions. However, systematic underperformance with respect to the null model was observed when only the O_3_ parameter was used. These results allowed us to build an informal logic approximation that accounts also for the unknown causative variables (Figure 3B). When the PM2.5 concentration is low, the capacity of the PM2.5 model to catch non escalating regions (true negatives) is high as few of them show outbreaks (SP>0.9). We thus introduced the unknown causative variables to explain the rising in the cases number when PM2.5 concentration is low (Figure 3B, left branch of the model). However, when the PM2.5 concentration is high, the O_3_ is contributing positively to the capacity of the model in predicting the outbreaks (Figure 4A, SE=0.64). This implies the existence of a variable threshold, in this case PM2.5 concentration, that acts on the O_3_ predictive capacity. This threshold is exemplified in the right branch of the model in Figure 3B.

SARS-CoV-2 is an emerging pathogen, and our study is with this regard subjected to possible limitations, primarily access to data and time constraints. Our model was validated based on publicly available data, different sampling policies and unknown fractions of untested/asymptomatic infected individuals, all of which could impact the accuracy of the collected datasets. Moreover, data access to the broader set of atmospheric factors with more powerful modeling strategies could be applied to establish explicit causal relation with the infection dynamic. Finally, the findings of our study should be seen in the light of the ongoing new pandemic, which makes our analysis short time windowed. On this point, temporal synchronization based on epidemiological parameters, was not implemented to synchronize the infection dynamics.

Nevertheless, our data support the concept that the atmospheric conditions can both 1) promote the formation of persisting forms of airborne droplets charged with SARS-CoV-2 (PM2.5), and 2) reduce the activity of the virus (O_3_). We thus hypothesized that the increase of the concentration of PM2.5 may reflect the rise of infective droplets with a diameter inferior to 5 microns. We speculate that fast evaporation of droplets emitted by talking or sneezing increases the sustained circulation of micro-droplets that carry a higher viral load, particularly in closed spaces.

In line with this, it has been shown for an airborne influenza virus that 49% of the viral particles are present in the droplets with a dimension between 1 and 4 μm. In general, droplets with a dimension bigger than 50 μm fall immediately to the ground, whereas particles with a dimension of 5 μm take more than an hour to reach the ground from a height of 3m (Tellier 2009). Moreover, particles with an aerodynamic diameter smaller than 2 μm remain suspended in the air for hours or days and are more able to reach the alveolar regions in the lungs (Shaman and Kohn 2009).

The droplet size can be modulated by the difference in vapour pressure, reducing it in a few seconds, at a rate of 5μm/sec (Wang et al. 2016). These observations are remarkably significant as they show that 1) the distribution of the dimension of the droplets changes immediately so that the diameter at the emission source (infected individual) is consistently bigger than the diameter at the destination (susceptible individual), and 2) the viral concentration in the droplet (viral particles/μl) increases, as the volume decreases quadratically respect to the radius. These evidence suggest that in unfavorable ambiental conditions, monitored by PM2.5 formation, the amount of persistent droplets with large viral load reflects on a significant increase of the hospitalized cases (Figure 5A). This supports the findings that face mask usage can be more beneficial in avoiding the spreading of the virus from an infected individual than to block the viral particles at the susceptible (healthy) individual (Leung et al. 2020).

Finally, the environmental factors that influence the particulate size/concentration, may be also responsible for the seasonal dynamics of the airborne infections. Our data are in agreement with this possibility (Figure 4C), however, a more detailed analysis is required to prove this causation for SARS-CoV-2 infection. To continue exploring seasonal and daily trends, the updated predictions in the 154 regions of our model with PM2.5 and O_3_ as input variables, can be found at https://github.com/COVID19Upcome/Europe.

In conclusion, our analysis supports the hypothesis that the atmospheric conditions that increase the particulate matter formation, are also contributing to the severity of the SARS-CoV-2 infection. An appealing possibility that ozone might act to counteract/sterilize viral charge is to be further investigated. Finally, monitoring spatial-temporal variations of atmospheric particulate and O_3_ could be used as an aid to estimate upcoming trends for the SARS-CoV-2 transmission impact.

## Data Availability

Manuscript is under evaluation in a peer review journal, and data will be available upon response from the editors.

## Supplementary figures

**Figure S1.**
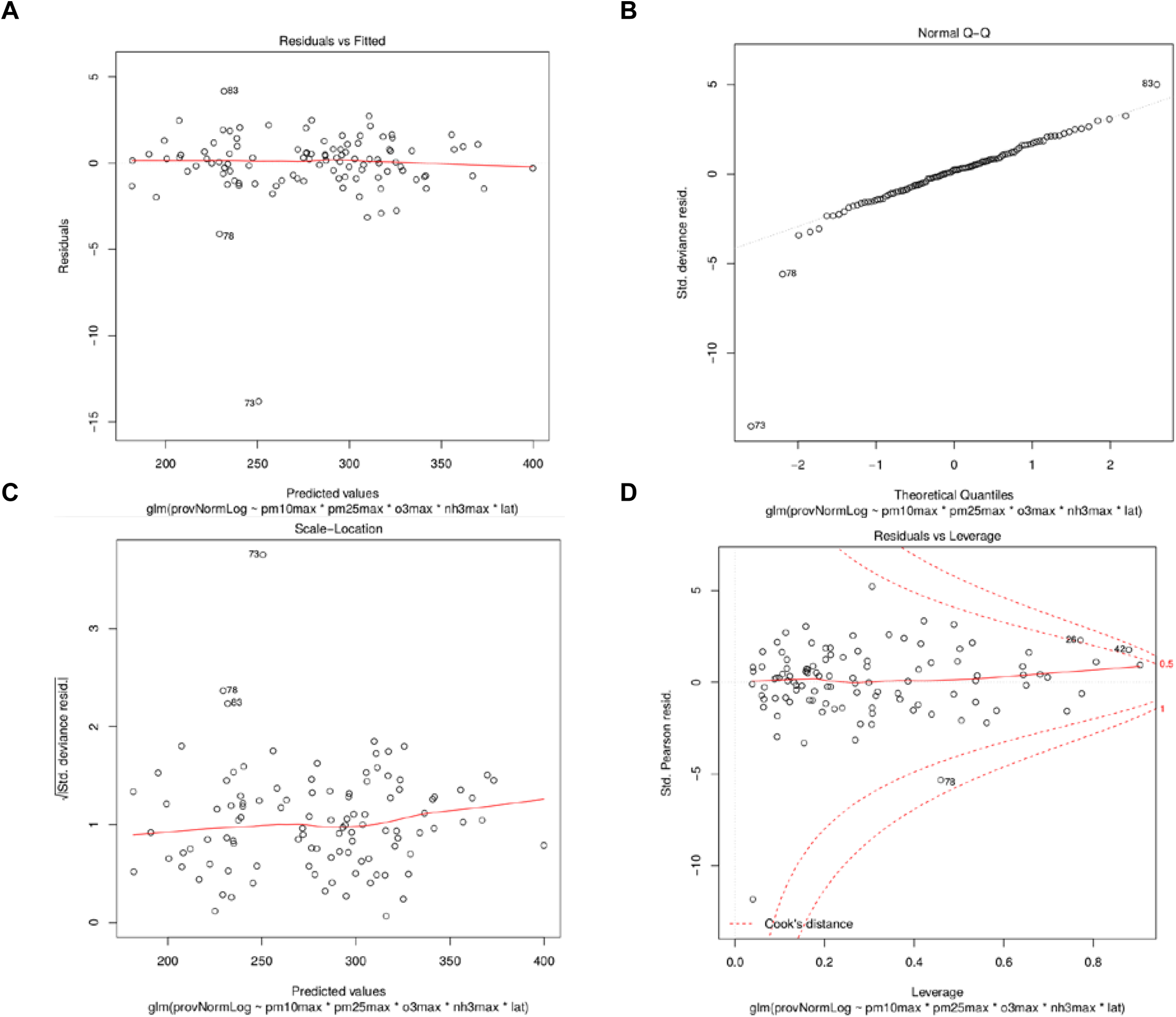
Evaluation of the Poisson model. The four diagnostic plots verify the quality of the full model in respect to the residuals (A), the non-normality errors (B), the homoscedasticity (C), and the influence of the outliers (D). The explaining variables considered where *AdM*_*PM*2.5_, *AdM_PM_*_10_, 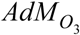, 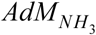, latitude and their combinations.

**Figure S2.**
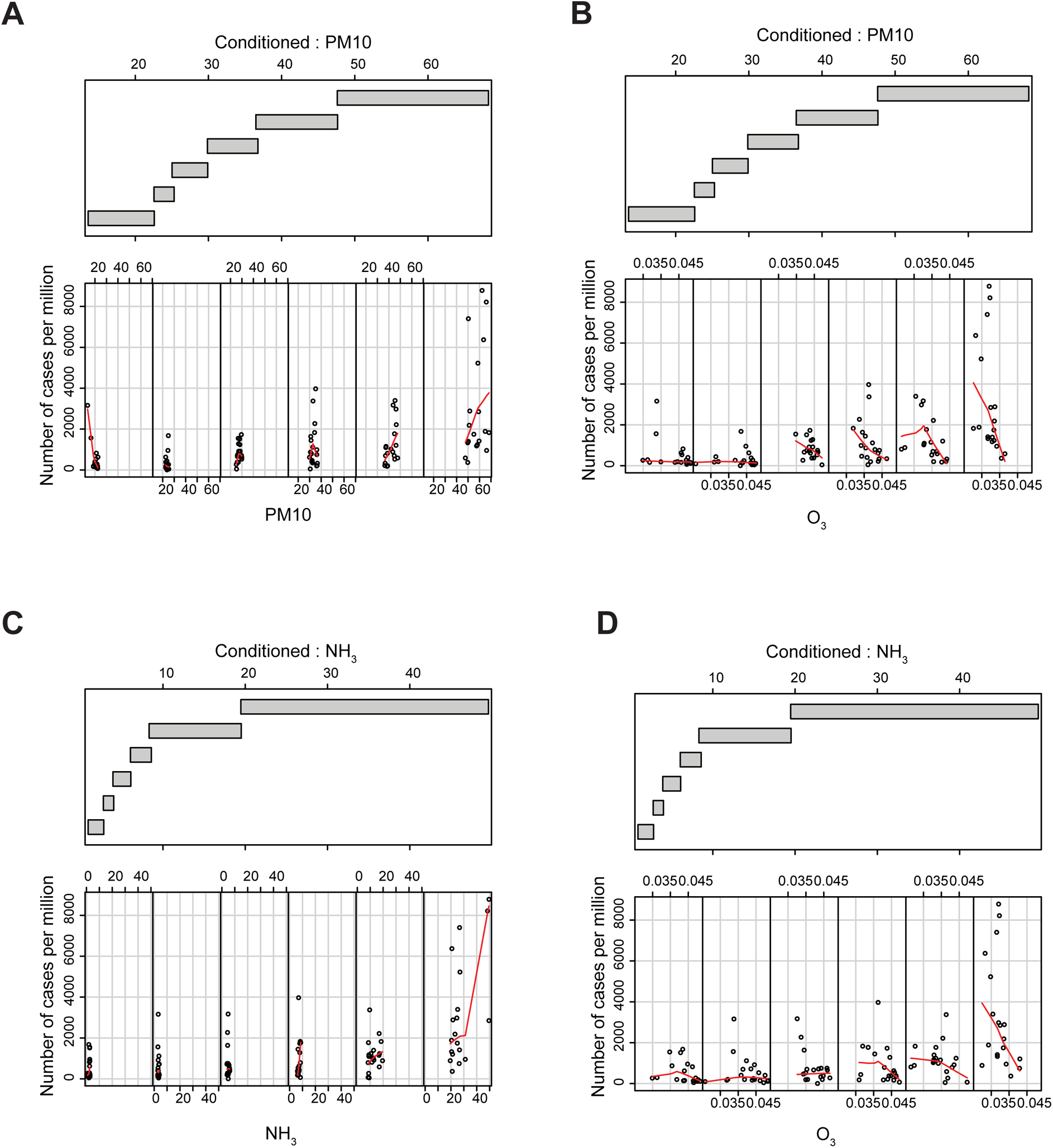
Evaluation of the cross-influence among PM10, NH_3_ and O_3_. Conditioned plots with six levels of conditioning. Lower panel: the scatter plots between the selected factor and the number of cases per million. Each plot is restricted to show data points that belong to the provinces that fall in the corresponding range of the conditioning factor. Upper plot: the range of the values that define each level of conditioning. The overlapping among the levels is 0.1. The lowess curve is computed in each scatterplot and shown in red. A) the scatterplot of PM10 conditioned to PM10; B) the scatterplot of O_3_ conditioned to PM10; C) the scatterplot of NH_3_ conditioned to NH_3_; D) the scatterplot of O_3_ conditioned to NH_3_

**Figure S3.**
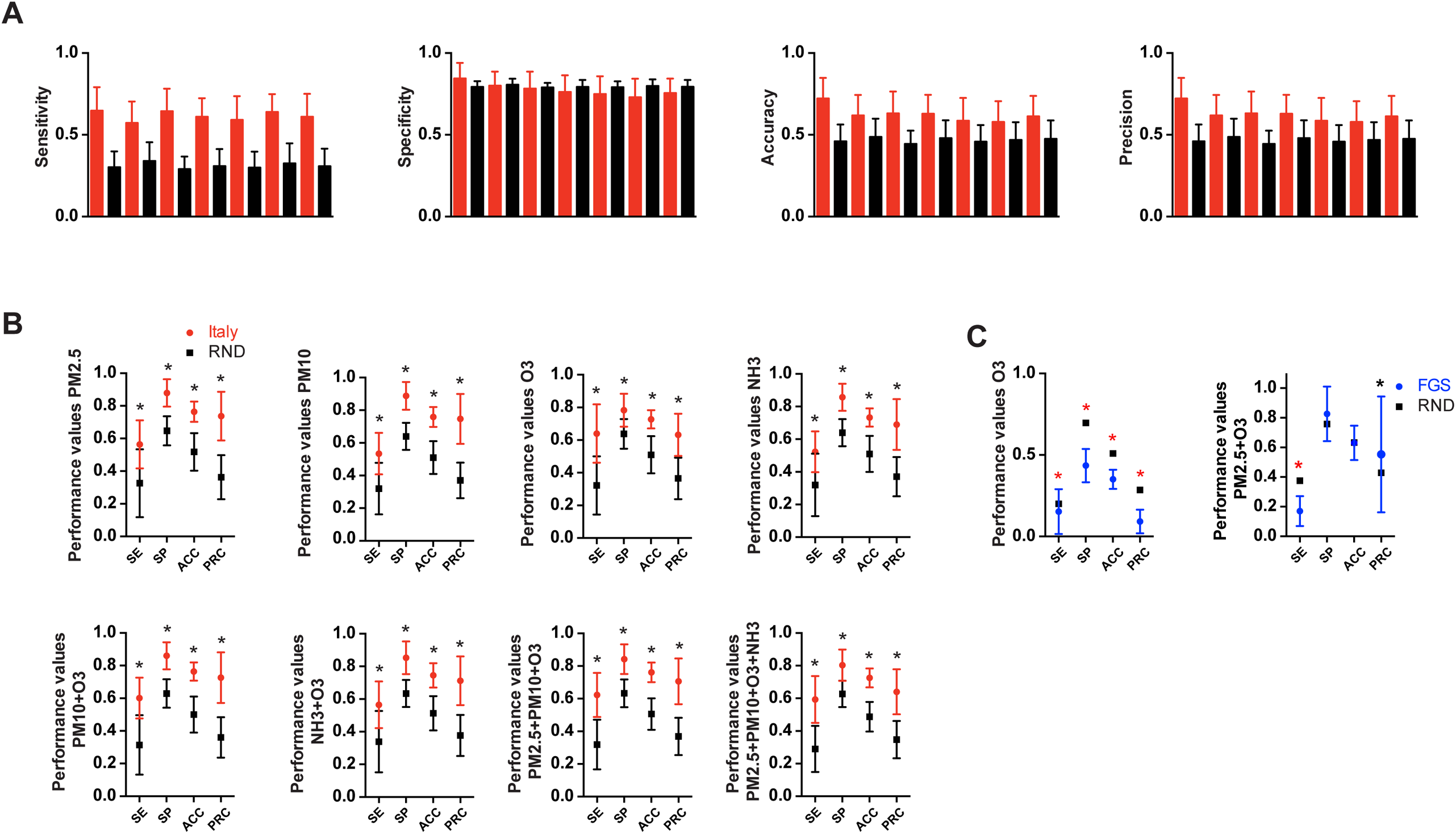
ANN performance evaluation. A) Performance indexes of the ANN varying the number of hidden nodes from 3 to 15 (step of 2). The bars represent the standard deviations of 100 Monte Carlo cross-validations. Italian provinces data are represented in red histograms, random data in black histograms. B) Performance values of the ANN on the 107 Italian provinces data varying the input variables. SE sensitivity, SP specificity, ACC accuracy, PRC precision. The dots represent the ANN average performance based on 100 Monte Carlo cross-validations. Bars represent the standard deviation. Italian provinces (red dots); France, Germany and Spain (FGS) regions (blue dots) and random dataset (black dots). C) Performance values of the ANN on the 47 FGS regions using a single atmospheric value (PM2.5, O_3_). FGS (blue dots), and Random dataset (black dots). Statistical analysis were performed using multiple t test corrected with Sidak-Boneferroni method for multiple comparisons (* p-value<0.001). Red asterisk indicates that the classifier performs worse than the null classifier.

## Acknowledgments

The authors thank Dr. Iva Lucic, Dr. Federico Colombo and Dr. Carlotta Penzo for proofreading and helpful suggestions.

## Author Contributions

R.F. and B.L. conceived the project. R.F. designed the experiments. R.F. performed in silico experiments and bioinformatic analysis. R.F. and B.L. analyzed data. R.F., M.L., M.S. and B.L. wrote the manuscript.

